# Quantifying the benefits of improved operations for polio outbreak response: A model-based analysis

**DOI:** 10.1101/2025.10.15.25338096

**Authors:** Yuming Sun, Pinar Keskinocak, Stephanie D. Kovacs, Lauren N. Steimle

## Abstract

**Background:** Novel type 2-containing oral poliovirus vaccine (nOPV2) was introduced in 2021. Since then, over 1 billion doses of nOPV2 have been used in outbreak response globally. Despite the reduced risk of virus reversion with nOPV2, stopping ongoing circulating vaccine-derived poliovirus type 2 (cVDPV2) outbreaks in some countries and achieving global cVDPV2 elimination have proved challenging, which highlights the potential needs to update outbreak response operations. In this study, we adapted and extended an existing poliovirus transmission model to evaluate the impact of two operational changes in outbreak response: prioritizing under-vaccinated individuals to receive nOPV2 in each vaccination round and increasing the number of vaccination rounds in each response.

**Methods:** We tested several outbreak response scenarios, assuming a limited nOPV2 stockpile. These scenarios varied in the strategy of allocating nOPV2 doses among target individuals of a response: (1) *Baseline allocation* where each target individual had equal chance of receiving nOPV2; (2) *Priority allocation* where “under-vaccinated individuals” were prioritized to receive nOPV2; Under-vaccinated individuals were identified based on the history of, either type 2-containing OPV doses received through outbreak response or inactivated poliovirus vaccine (IPV) doses received through essential immunization; (3) *Idealized allocation* where “low-immunity individuals” were prioritized to receive nOPV2; Low-immunity individuals were identified based on true immunity level. These scenarios also varied in the number of vaccination rounds (i.e., 3 to 5 rounds) in the response to each detection of cVDPV2 transmission. Outcome measures included outbreak size (i.e., number of cVDPV2 paralytic cases), time to outbreak interruption, and total nOPV2 doses used in outbreak response. We tested these scenarios by predicting cVDPV2 transmission in Nigeria for the study period of 2024-2028.

**Results:** Interruption of cVDPV2 transmission in Nigeria was only achieved with idealized allocation and 5 vaccination rounds in each response. In all other scenarios, cVDPV2 transmission continued at the end of the study period. Compared to the baseline allocation, with the same number of vaccination rounds, priority allocation based on the history of type 2-containing OPV doses received through outbreak response resulted in smaller outbreak sizes using fewer nOPV2 doses; priority allocation based on the history of IPV doses received through essential immunization resulted in larger outbreak sizes using more nOPV2 doses.

**Conclusions:** Prioritizing under-vaccinated individuals based on type 2-containing OPV vaccination history along with increasing the number of vaccination rounds can reduce cVDPV2 transmission. However, reactive use of nOPV2 in outbreak response may not suffice to eliminate cVDPV2 outbreaks in high-risk countries such as Nigeria, as the required operational changes (e.g., prioritization of low-immunity individuals and 5 rounds) might supersede the response capacity in these countries. Incorporating nOPV2 into essential immunization or other preventive vaccinations need to be considered to stop cVDPV2 transmission in high-risk countries and achieve global cVDPV2 elimination.

## 1. Introduction

The Global Polio Eradication Initiative (GPEI) launched in 1988 [1] has made remarkable progress in reducing poliovirus transmission through the use of oral poliovirus vaccine (OPV) in essential immunization and vaccination campaigns. Wild poliovirus (WPV) types 2 and 3 were certified as eradicated in 2015 [2] and 2019 [3], respectively. WPV type 1 remains endemic only in Pakistan and Afghanistan. Overall, GPEI efforts have reduced global polio cases by more than 99% and prevented millions of cases of paralytic polio.

Despite these achievements, circulating vaccine-derived poliovirus (cVDPV) has emerged as a significant challenge to eradication. cVDPV occurs when the live-attenuated virus contained in OPV genetically reverts and regains transmissibility and neurovirulence, particularly in populations with insufficient immunity to halt virus circulation. To mitigate this risk, GPEI adopted a phased cessation of OPV use by serotype, following the eradication of the homotypic WPV. The first step was the global cessation of type 2-containing OPV (OPV2) in 2016. It was implemented through a globally coordinated “switch” from trivalent OPV (tOPV; containing all three serotypes) to bivalent OPV (bOPV; containing serotypes 1 and 3) in essential immunization that aims to vaccinate newborns at specific ages (e.g., 6, 10, and 14 weeks old [4]). To maintain immunity against type 2 poliovirus after OPV2 cessation, in 2015 before the switch, the GPEI supported the introduction of one dose of inactivated poliovirus vaccine (IPV; containing all three serotypes) in all countries using OPV in essential immunization [5]. Since 2020, because of the increased emergence and transmission of cVDPV type 2 (cVDPV2), the GPEI recommended adding a second IPV dose [6]. Because IPV contains inactivated poliovirus, its use does not result in cVDPV emergence. IPV provides strong protection against paralysis once infected by the poliovirus. However, it has limited protection against infection and transmission through the fecal-oral route in individuals not previously exposed to live poliovirus (e.g., WPV, cVDPV, and the virus contained in OPV). This property of IPV makes it less effective to stop outbreaks in settings with suboptimal hygiene and sanitation and high force of transmission.

Since the switch in 2016, cVDPV2 outbreaks have accounted for over 60% of global polio cases [7]. These outbreaks were driven by several factors, including the failure to eliminate existing cVDPV2 before the switch, insufficient coverage with IPV in essential immunization, and the limited ability of IPV to interrupt transmission [8, 9]. In addition, vaccination campaigns using monovalent OPV2 (mOPV2) in response to cVDPV2 outbreaks were delayed, and they had insufficient scope and coverage to interrupt existing transmission and prevent seeding new cVDPV2 emergences [8, 9]. Vaccination campaigns conducted during outbreak response usually target children of a broader age group (e.g., 0-4 years old) over a short period (e.g., 4 days). Each response involves multiple rounds of vaccination campaigns, also known as outbreak response supplementary immunization activities (oSIAs).

To mitigate the risk of seeding new cVDPV2 from using mOPV2 in post-switch outbreak response, a novel OPV2 (nOPV2) was developed with enhanced genetic stability and reduced risk of reversion [10]. However, despite the deployment of over one billion nOPV2 doses globally since 2021 [11], cVDPV2 outbreaks continue in over 30 countries as of 2025 [12]. These persistent outbreaks suggest that the core problem is not OPV itself but the operational performance of outbreak responses. Stakeholders are raising questions about whether relying on outbreak response using nOPV2 can successfully interrupt cVDPV2 outbreaks, and if so, what operational improvements are needed to achieve this goal.

Previous modeling studies suggested that stopping cVDPV2 outbreaks required improvements in multiple aspects of outbreak response, such as its timeliness and coverage, particularly in under-vaccinated areas [13-18]. Some studies suggested that, during outbreak response, stakeholders should prioritize the vaccinations of “low-immunity individuals” caused by, e.g., missed vaccinations, vaccine failure, or absence of prior live poliovirus infections, especially when vaccine supply is limited [18]. However, implementing these improvements is challenging in most of the areas affected by cVDPV2. Improving timeliness of outbreak response is often not feasible because of logistical constraints in mobilizing vaccinators and obtaining vaccines and other supplies. Achieving high coverage in areas isolated by conflict, remoteness, or social marginalization is resource-intensive and difficult to monitor. Identifying low-immunity individuals before a campaign is also impractical, as no rapid and accurate immunity tests are currently available.

Given these challenges, we considered two practical operational improvements to outbreak response: increasing the number of oSIA rounds in each response and prioritizing “under-vaccinated individuals” to receive nOPV2 in each campaign. We referred to under-vaccinated individuals as those with few or no recorded poliovirus vaccine doses in their available vaccination histories, regardless of their true immunity levels. The aim of this study was to answer whether the aforementioned operational improvements of outbreak response could reduce the prevalence of polio cases, improve the efficient use of the limited nOPV2 stockpile, and finally, successfully interrupt persistent cVDPV2 transmission.

We evaluated numerous outbreak response scenarios, using Nigeria’s cVDPV2 transmission as a case study. These scenarios varied from 3 to 5 oSIA rounds conducted in the response to each detection of transmission and varied how a limited supply of nOPV2 doses was allocated among target individuals in each round. Tested vaccine allocation strategies during an oSIA round included: (1) a *baseline allocation* in which no one was prioritized and thus all target individuals had an equal chance of receiving nOPV2; (2) a *priority allocation*, that prioritized nOPV2 doses to under-vaccinated individuals based on the history of IPV doses received through essential immunization (2a) or OPV2 doses received through oSIAs (2b); and (3) an *idealized allocation* that prioritized nOPV2 doses to low-immunity individuals based on true immunity level. In all scenarios, we assumed adherence to the standard operating procedures regarding coverage, target age groups, timeliness, and the interval between consecutive rounds [19]. These included covering 90% of children aged 0-4 years in each round’s target geographic area and initiating the first round 4 weeks after a detection of transmission, with subsequent rounds spaced at 4-week intervals.

## 2. Material and methods

### 2.1. Live poliovirus transmission model

#### 2.1.1. Model compartments

To enable the identification of under-vaccinated individuals, we extended a previously developed live poliovirus transmission model [16, 17]. The model is general and can be applied to simulate the transmission of one serotype at a time. The population in the model is divided into four compartments: Susceptible (*S*) – at risk of infection or can be vaccinated; Exposed (*E*) – infected by live poliovirus (including those who received an “effective” OPV dose) but not yet infectious to others; Infectious (*I*) – infected by live poliovirus (including those who received an effective OPV dose) and can transmit the virus to others; and IPV-reacted (*H*) – recently received an effective IPV dose but has not acquired the corresponding immunity induced by this dose yet. An effective vaccine dose refers to a dose that successfully elicits immune response in the recipient which thereafter provides a measurable level of immunity. Each of these compartments is further stratified according to five factors (with their corresponding subscripts), including immunity group (*i*), vaccination group (*k*), virus strain (*j*), age group (*a*), and subpopulation (*s*).

Eight immunity groups are considered based on the source of immunity, the timing of the most recent immunizing event, the number of exposures to live poliovirus, and the number of effective IPV doses. Immunity groups are ordered by increasing immunity levels from 0 (least immunized) to 7 (most immunized). They include unimmunized individuals with no immunity against poliovirus (immunity group 0), IPV-only-immunized individuals who only have humoral immunity (immunity groups 1-4), and live-poliovirus-immunized individuals who have intestinal mucosal immunity (immunity groups 5-7) (see Supplemental Materials S1).

The model has eight vaccination groups from 0 (least vaccinated) to 7 (most vaccinated). These include unvaccinated individuals with no poliovirus vaccines (vaccination group 0), IPV-only-vaccinated individuals who have received only IPV (vaccination groups 1-4), and OPV-vaccinated individuals who have received OPV (vaccination groups 5-7) (see Supplemental Materials S1). Unlike immunity groups that reflect actual immunity levels, vaccination groups only reflect individuals’ recorded vaccination histories on the number and dates of poliovirus vaccine doses received through previous vaccinations, regardless of whether these doses are effective or not. For example, a child who just received their first IPV dose at 6 weeks of age according to the vaccination card has a vaccination history of “1 IPV dose at 6 weeks” and thus is included in vaccination group 2. However, this child might remain unimmunized and stay in immunity group 0 if this IPV dose is not effective.

Twenty hypothetical virus strains are included to capture the reversion progress of the live-attenuated virus contained in OPV. Depending on which OPV formulation is being simulated, strain 1 represents either the genetically stabilized strain contained in nOPV or the Sabin strain contained in Sabin-strain OPV (tOPV, bOPV, and mOPV), and strains 2-19 represent the partially reverted forms of the genetically stabilized strain or the Sabin strain. Strain 20 represents cVDPV. Each virus strain is characterized by the basic reproductive number for transmissibility and by the paralysis-to-infection rate for neurovirulence. Compared to the Sabin strain, the model assumes a reduced but non-zero reversion of the genetically stabilized strain (see Supplemental Materials S1).

Age groups are modeled to enable preferential mixing among similar ages, the age-dependent essential immunization schedule, and the potential transmission in all ages. The model also considers non-overlapping subpopulations based on geography, historical vaccination coverage, and accessibility. The specific characterizations of age groups and subpopulations depend on the studied population, with more details in [16, 17].

Altogether, the model consists of the following compartments:

- *S*_*i,k,a,s*_: Susceptible individuals in immunity group *i*, vaccination group *k*, age group *a*, and subpopulation *s*;
- *E*_*i,k,j,a,s*_: Exposed individuals infected by virus strain *j*, in immunity group *i*, vaccination group *k*, age group *a*, and subpopulation *s*;
- *I*_*i,k,j,a,s*_: Infectious individuals infected by virus strain *j*, in immunity group *i*, vaccination group *k*, age group *a*, and subpopulation *s*;
- *H*_*i,k,a,s*_: IPV-reacted individuals in immunity group *i*, vaccination group *k*, age group *a*, and subpopulation *s*.

#### 2.1.2. Transitions between model compartments

Individuals transition between model compartments as a result of birth, death, aging, disease dynamics (e.g., recovery from infection), OPV reversion, waning immunity, infection, and vaccination. In this study, we updated the transitions caused by vaccination. Vaccination changes an individual’s immunity group, if the vaccine dose received is effective, and vaccination group, if the vaccine dose is recorded in their vaccination history. Whether a vaccine dose is effective is determined by the estimated per-dose vaccine effectiveness between 0 and 1 (see 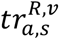and 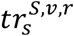in Supplemental Materials S1). Whether a vaccine dose is recorded is based on the pre-defined recording probability between 0 and 1 (see 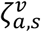and 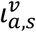in Supplemental Materials S1).

Figure 1 provides a simplified illustration on all possible transitions of a susceptible individual in compartment *S*_*i,k,a,s*_ after getting an IPV dose, an OPV dose, and an infection of virus strain *j*, while more details on transitions between model compartments are available in [16, 17].

**Figure 1:**
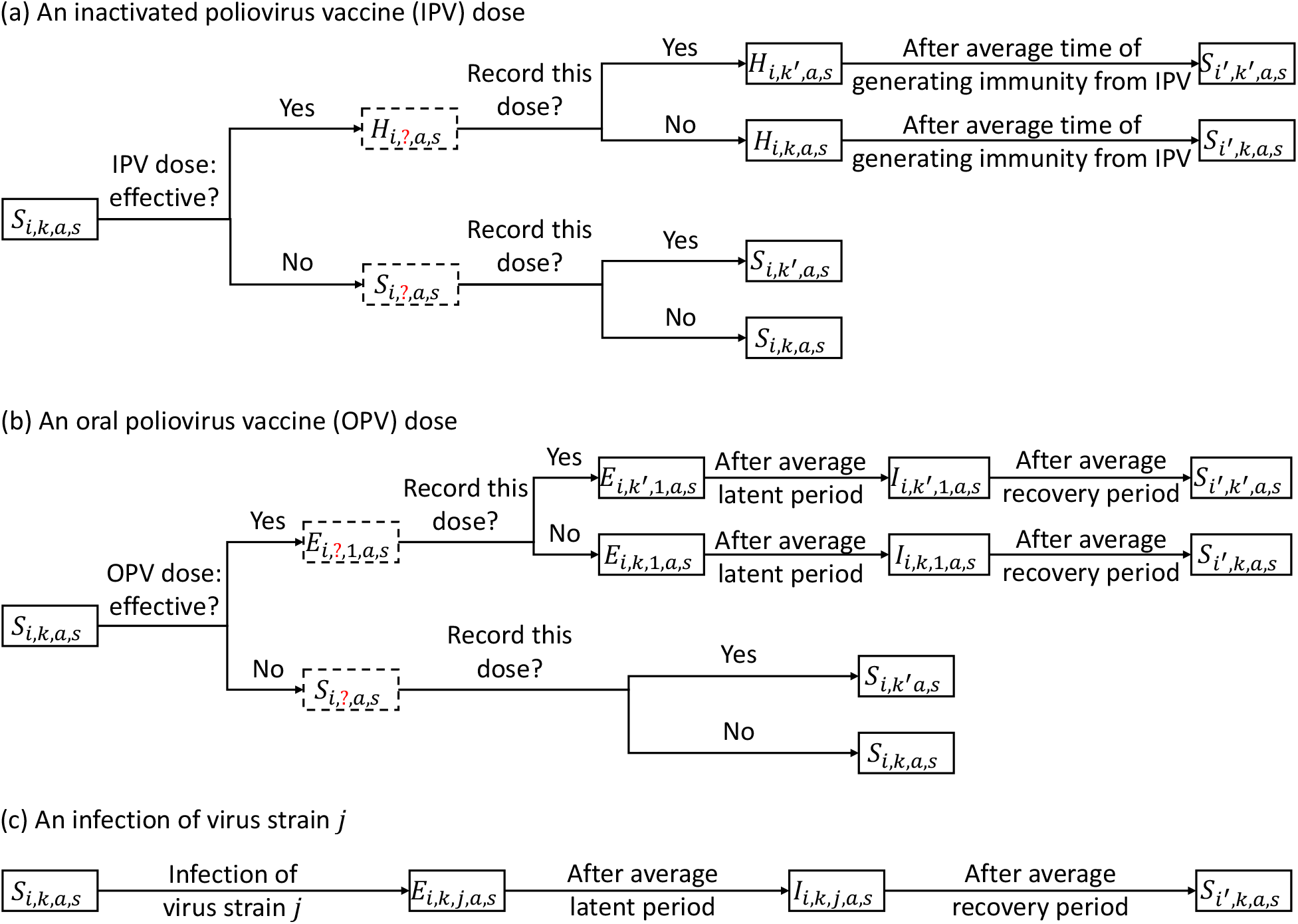
Simplified illustration on transitions between model compartments caused by getting (a) an inactivated poliovirus vaccine (IPV) dose; (b) an oral poliovirus vaccine (OPV) dose; and an infection of virus strain *j*. Dashed boxes indicate an intermediate transition.

##### (1) Transitions after getting an IPV dose

As shown in Figure 1(a), if the IPV dose is effective, the individual will transition to a IPV-reacted compartment, *H*, for a short period while they mount the immune response. If this effective dose is recorded in their vaccination history, the individual’s vaccination group will change from *k* to *k*^′^, i.e., they will transition to *H*_*i,k*′,*a,s*_, otherwise, *H*_*i,k,a,s*_. After developing the immunity induced by this effective IPV dose, the individual will transition to a susceptible compartment of an updated immunity group *i*^′^, i.e., *S*_*i*′,*k*′,*a,s*_ when the IPV dose is recorded and *S*_*i*′,*k,a,s*_ otherwise.

If the IPV dose is not effective, the individual will remain in a susceptible compartment *S* because no immunity will be induced. If the dose is recorded, the vaccination group will be updated from *k* to *k*^′^, and the transition will be to *S*_*i,k*′,*a,s*_; otherwise, the individual will stay in *S*_*i,k,a,s*_.

Feasible changes from vaccination group *k* (immunity group *i*) to vaccination group *k*^′^ (immunity group *i*^′^) depend on *k* (*i*) and whether IPV or OPV is received. For example, if this individual in compartment *S*_*i,k,a,s*_ is unvaccinated (*k* = 0) and unimmunized (*i* = 0), after receiving and recording an IPV dose, their vaccination group will change to *k*^′^ = 2 (IPV-only-vaccinated who received the most recent IPV dose within the past two years and had only 1 IPV dose in life). If it is an effective IPV dose, their immunity group will change to *i*^′^ = 2 (IPV-only-immunized who received the most recent effective IPV dose within the past two years and had only 1 effective IPV dose in life). See Tables S1 and S2 in Supplemental Materials S1 for all combinations of *k* and *k*^′^ and of *i* and *i*^′^.

##### (2) Transitions after getting an OPV dose

Possible transitions for an individual in compartment *S*_*i,k,a,s*_ after receiving an OPV dose are similar to that after an IPV dose (see Figure 1(b)). Their vaccination group will change if the OPV dose is recorded and their immunity group will change if the OPV dose is effective. However, this individual will transition to an exposed compartment *E* of virus strain 1 (representing the live-attenuated virus contained in OPV) instead of an IPV-reacted compartment *H*, then to an infectious compartment *I* of virus strain 1, and finally to a susceptible compartment *S* after the average recovery period. Since the live-attenuated virus contained in OPV is live poliovirus, the model treats the transitions caused by OPV vaccination the same as that of live poliovirus infection, enabling the effect of passive immunization [20].

##### (3) Transitions after getting a live poliovirus infection

The infection of live poliovirus changes an individual’s immunity group, but not their vaccination group, since infections are not recorded in their vaccination history (see Figure 1(c)).

### 2.2. Simulations of cVDPV2 outbreaks in Nigeria

#### 2.2.1. Model validation and parameters

We adapted model parameters from a case study of cVDPV2 outbreaks in the Northwest and Northeast geopolitical zones of Nigeria [16-18]. Most of the parameter values were estimated from existing polio models and data on epidemiology and demography (e.g., vaccine effectiveness and vaccination coverage in different geographic areas). Some uncertain parameters such as the mixing between subpopulations were estimated through iterative model calibration and validation [16, 17].

We characterized eleven age groups, ages 0-2 and 3-11 months and ages 1, 2, 3, 4, 5-9, 10-14, 15-24, 25-39 and ≥ 40 years. We set up five non-isolated subpopulations which had some accessibility for population movement, surveillance and vaccinations (see Figure 2). Two subpopulations were considered *general* given higher historic vaccination coverage and 100% accessibility and were located in Northwest Nigeria (subpopulation 1: Jigawa, Kaduna, Kano, and Katsina) and Northeast Nigeria (subpopulation 3: Adamawa, Gombe, and Taraba). Three subpopulations were considered *under-vaccinated* because of lower vaccination coverage and/or accessibility. They included subpopulation 2 in Northwest Nigeria (Kebbi, Sokoto, and Zamfara) and subpopulations 4 (Bauchi, Borno, and Yobe) and 5 (areas in Borno and Yobe with insurgency and lower accessibility) in Northeast Nigeria. The other two isolated subpopulations (local government areas Abadam and Marte in Borno) from previous studies [16-18] were not included here, because we assumed they had 0% accessibility for security reasons and no poliovirus transmission.

**Figure 2:**
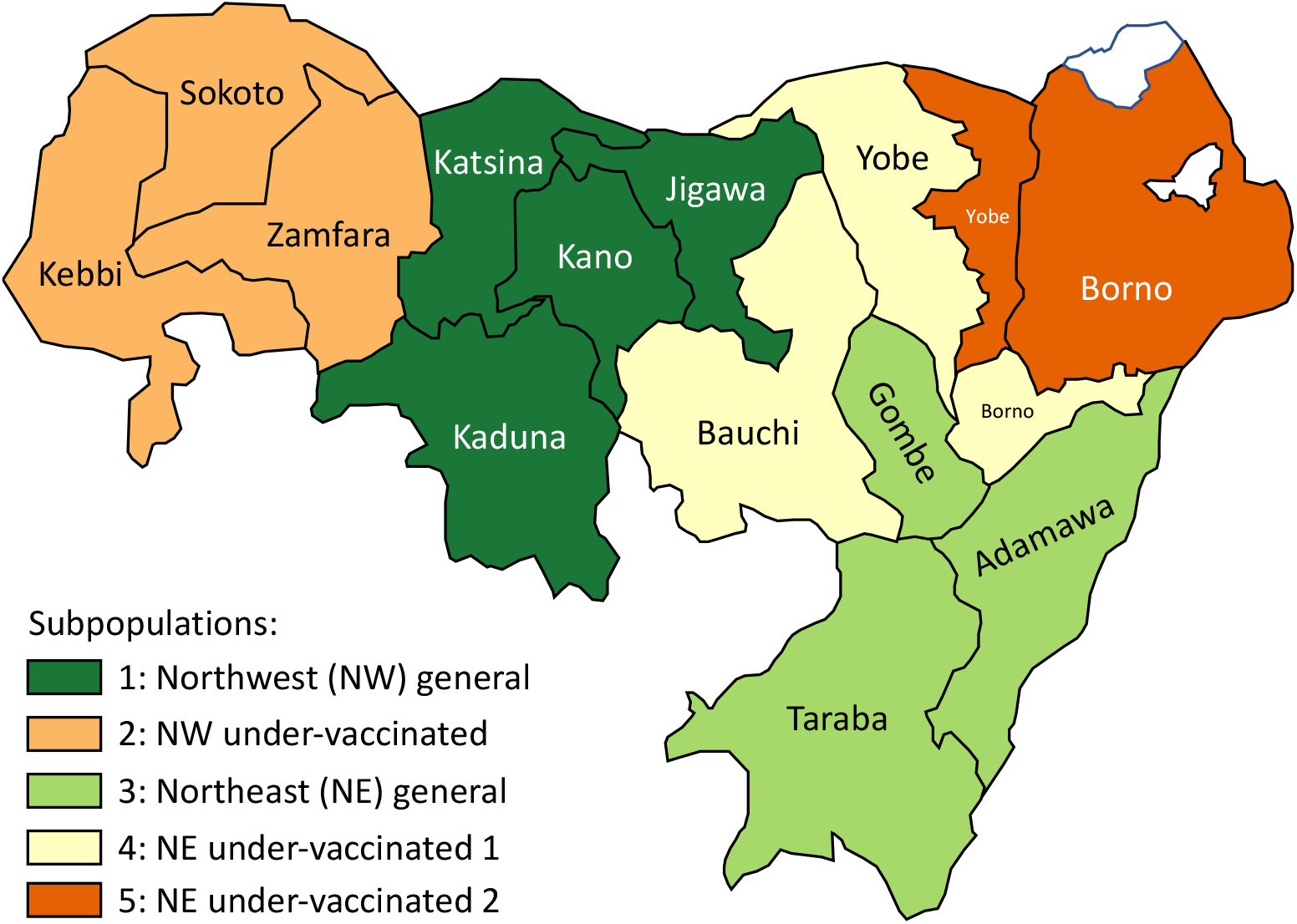
Non-isolated subpopulations in Northwest and Northeast Nigeria modeled.

To seed the simulations, we updated the “initial condition” in the previous Nigeria study [18] to incorporate individuals of different vaccination groups. The initial condition specified how many individuals were in each model compartment at the start of simulations, i.e., 2024. We estimated the distribution of the population across vaccination groups, using available information on individual-level vaccination records. Usually, individuals’ vaccination cards record the date of administration of each vaccine dose delivered through essential immunization. Data from vaccination cards are aggregated to estimate essential immunization coverage in each birth cohort at the district, provincial, and national levels [21]. Nigeria’s essential immunization program co-administered IPV with the Diphtheria, Tetanus, and Pertussis (DTP) vaccine, using a schedule of one IPV dose at 14 weeks during 2015-2021, and two IPV doses at 6 and 14 weeks, respectively, since 2021. We estimated essential immunization coverage with one IPV dose (i.e., IPV1) and two IPV doses (i.e., IPV2) among children aged 0-4 years at the start of 2024, aggregating IPV coverage data from Nigeria’s Demographic and Health Survey 2023-2024 [22] and DTP coverage data of 2019-2024 from the Institute for Health Metrics and Evaluation [23]. We then used the estimated IPV1 and IPV2 to determine the number of children aged 0-4 years in vaccination group 0 (unvaccinated) and vaccination groups 1-4 (IPV-only-vaccinated) at the start of 2024. In the initial condition, we did not estimate the numbers of children aged 0-4 years in those OPV-vaccinated groups 5-7, because no type 2-containing OPV doses were delivered through essential immunization and individual-level records of type 2-containing OPV received through outbreak response are not available. Also, we assumed all individuals aged ≥ 5 years were in vaccination group 0 at the start of 2024. The distribution of them across vaccination groups does not impact the transmission dynamics when oSIAs target only those under 5 years old. See more details of estimating the initial condition in Supplemental Materials S2.

Since the model considers a reduced reversion for the genetically stabilized strain contained in nOPV compared to the Sabin strain, we assumed this reduction would also result in a lower vaccine effectiveness. Therefore, we set the effectiveness of nOPV2 to 90% of that of monovalent OPV2.

In the model, a detection of poliovirus transmission in a subpopulation occurs when the weekly number of new paralytic cases exceeds a pre-specified detection threshold of 1 case per week [18] in that subpopulation. Each detection of a poliovirus outbreak then triggers an outbreak response consisting of multiple oSIA rounds in that subpopulation.

We assumed the probability of a vaccine dose being recorded in an individual’s vaccination history and therefore changing their vaccination group was either 0 or 1, depending on the vaccine allocation strategy tested (see Section 2.2.2).

#### 2.2.2. Tested outbreak response scenarios in Nigeria

Using the validated model, we simulated numerous outbreak response scenarios over a study period of 2024-2028. We systematically varied two factors in an outbreak response: the number of oSIA rounds and the vaccine allocation strategy.

We tested the implementation of 3, 4, and 5 oSIA rounds in an outbreak response. Based on the standard operating procedures [19], we assumed that the first round would be implemented four weeks after a detection of transmission, followed by additional rounds every four weeks, with each round achieving 90% coverage. Each round targeted children aged 0–4 years, with a single dose of nOPV2.

The vaccine allocation strategy specified how limited nOPV2 doses in an oSIA round were allocated across target individuals, using different information on their vaccination needs (e.g., true immunity and vaccination history). In this study, we considered four vaccine allocation strategies, as summarized in Table 1.

**Table 1:** Tested vaccine allocation strategies in outbreak response scenarios.

| Notations | Vaccine allocation strategies |
| --- | --- |
| 1. $A^P$ | Baseline allocation |
| 2a. $A^V$-EI | Priority allocation based on the history of IPV received through essential immunization |
| 2b. $A^V$-oSIA | Priority allocation based on the history of OPV2 received through outbreak response supplementary immunization activities |
| 3. $A^M$ | Idealized allocation based on true immunity level |

We considered a *baseline allocation* strategy, *A*^*P*^, in which each target individual had an equal chance of receiving an nOPV2 dose. More specifically, under *A*^*P*^, the number of nOPV2 doses allocated to each model compartment was proportional to the number of individuals in that compartment. This strategy reflects current practice, in which oSIAs are conducted without consideration of individuals’ vaccination histories or immunity levels.

We considered two *priority allocation* strategies where under-vaccinated individuals (those in low vaccination groups) were prioritized to receive nOPV2 doses, based on the history of either IPV received through essential immunization (*A*^*V*^-EI) or OPV2 received through oSIAs (*A*^*V*^-oSIA).

As a comparison point, we also considered an *idealized allocation, A*^*M*^, where low-immunity individuals (those in low immunity groups) were prioritized to receive nOPV2 doses. We referred to *A*^*M*^ as the idealized allocation because individuals’ true immunity levels are not directly observable in practice as there is no quick and accurate polio immunity test. Compared to *A*^*M*^, *A*^*V*^-EI and *A*^*V*^-oSIA are more feasible to implement because individual’s vaccination history is more likely to be available.

Figure 3 illustrates how *A*^*P*^, *A*^*V*^ and *A*^*M*^ allocated limited nOPV2 doses across individuals differently through a simplified example. This example assumed three immunity groups, three vaccination groups, one age group, one subpopulation, and 12 susceptible individuals targeted by an oSIA round achieving a 50% coverage with 6 nOPV2 doses. Of notice, some individuals in a high vaccination group because of a history of multiple vaccine doses may have low immunity, if most of those doses were ineffective. Conversely, some individuals in the low vaccination groups may belong to a high immunity group if they have experienced infections by the live poliovirus. Under *A*^*P*^, 50% of the individuals in each compartment received nOPV2 (see Figure 3(b)). Under *A*^*V*^, nOPV2 allocation follows the priority order: vaccination group 0, followed by 1 and 2; thus, all 6 doses were used for the 6 individuals in vaccination group 0 (see Figure 3(c)). Under *A*^*M*^, nOPV2 allocation follows the priority order: immunity group 0, followed by 1 and 2; thus, all 6 doses were used for the 6 individuals in immunity group 0 (see Figure 3(d)).

**Figure 3:**
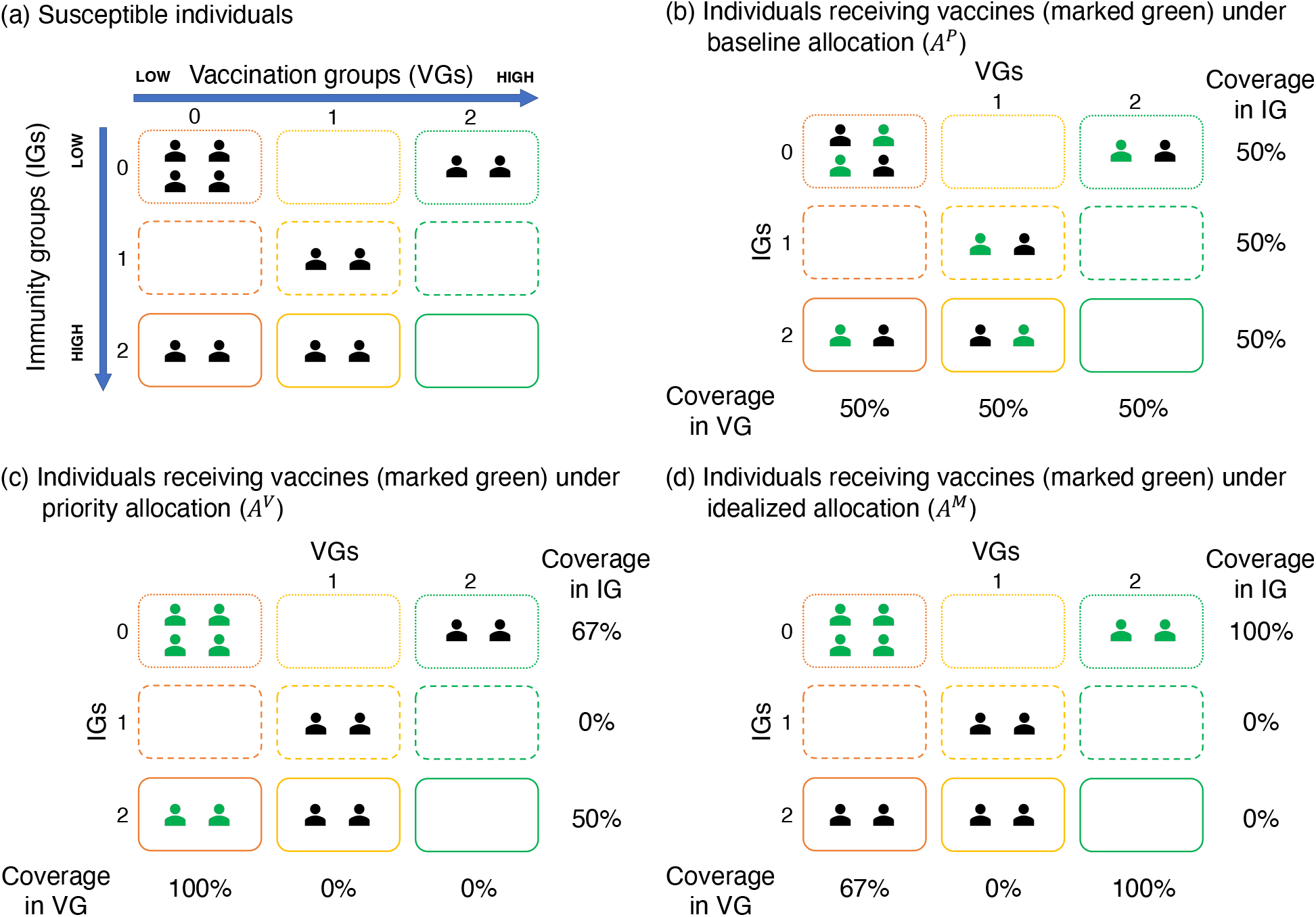
Simplified example on the differences between vaccine allocation strategies *A*^*P*^ (proportional allocation), *A*^*V*^ (priority allocation based on vaccination history), and *A*^*M*^ (priority allocation based on true immunity level)

These vaccine allocation strategies also differed in the changes in vaccination groups that the model tracked and, thus, the probability of a vaccine dose being recorded, despite all simulations starting with the same initial condition described in Section 2.2.1. When simulating *A*^*V*^-EI, the model tracked only changes caused by IPV doses received through essential immunization, setting the recording probability of an IPV dose delivered through essential immunization to 1 but that of all other doses to 0. Under *A*^*V*^-oSIA, the model tracked only changes caused by nOPV2 doses received through oSIAs, setting the recording probability of an nOPV2 dose delivered through oSIAs to 1 but that of all other doses to 0. Under *A*^*P*^ and *A*^*M*^, the model tracked no changes in vaccination groups, setting a recording probability of 0 for all doses for simplicity. Changes in vaccination groups do not impact the outcomes of *A*^*P*^ and *A*^*M*^, since neither strategy allocates vaccines based on vaccination groups.

#### 2.2.3. Outcome measures

We compared the outcomes of all scenarios by three measures: “time until die-out,” “outbreak size,*”* and “total nOPV2 doses administered.” Time until die-out measured the time point when the complete interruption of cVDPV2 transmission happened in the overall population (i.e., no more cVDPV2 infections in any subpopulations). The time-point was selected to be the first week in the study period when all subpopulations simultaneously had zero new cVDPV2 paralytic cases. Outbreak size evaluated the cumulative number of cVDPV2 paralytic cases over the study period. Total nOPV2 doses administered counted the cumulative number of nOPV2 doses administered through oSIAs over the study period. More details are in [16-18].

## 3. Results

This section reports all outcome measures after simulating the scenarios combining the number of oSIA rounds used in each outbreak response (3, 4 or 5 rounds) and the vaccine allocation strategy (*A*^*P*^, *A*^*V*^-EI, *A*^*V*^-oSIA, or *A*^*M*^).

Table 2 summarizes whether the interruption of cVDPV2 transmission was achieved in each scenario and the time until die-out in the scenarios where the transmission was interrupted. In our simulations, elimination was only achieved in one scenario, i.e., the idealized allocation based on true immunity level (*A*^*M*^) with 5 oSIA rounds in each outbreak response. This scenario’s time until die-out was 260 weeks, just before the end of the study period. In all other scenarios, cVDPV2 transmission continued in all subpopulations at the end of the study period, and therefore, time until die-out was undefined.

**Table 2:**
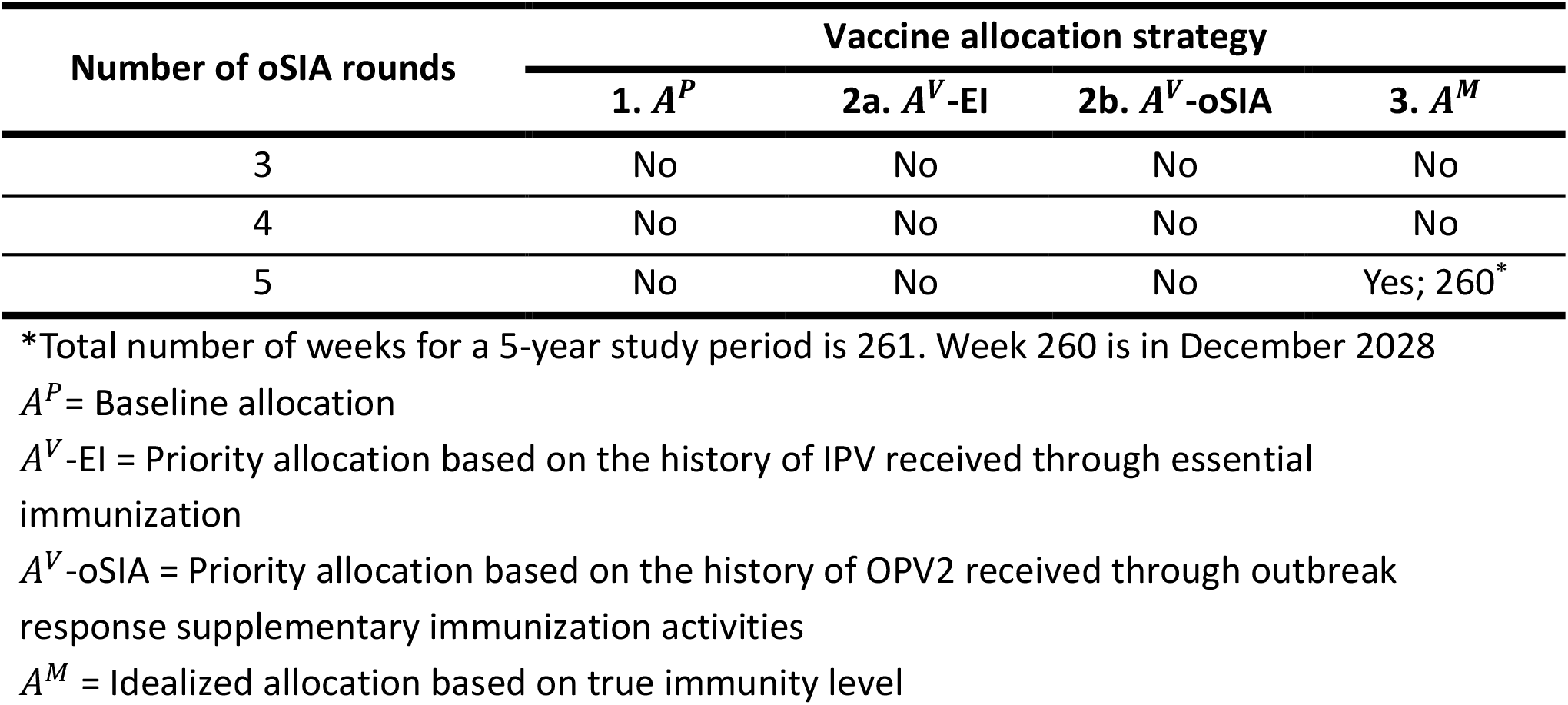
Simulation results for whether the interruption of cVDPV2 transmission was achieved (yes/no) and the time until die-out (in weeks) in tested outbreak response scenarios.

Table 3 and Figure 4 compare the outbreak size in number of paralytic cases and the total number of nOPV2 doses administered in all scenarios. We observed that, for any number of oSIA rounds used in an outbreak response, the *A*^*M*^ strategy with idealized vaccine allocation resulted in the smallest outbreak size (241-300 case counts) and required the fewest number of nOPV2 doses to deliver in oSIAs (204.4 to 248.1 million doses).

**Table 3:** Simulation results of outbreak size and total novel type 2-containing oral poliovirus vaccine (nOPV2) doses administered in tested outbreak response scenarios.

| Vaccine allocation strategies | Outbreak size<br>(in paralytic case counts) |  |  | Total nOPV2 doses administered<br>(in millions) |  |  |
| --- | --- | --- | --- | --- | --- | --- |
|  | 3 rounds | 4 rounds | 5 rounds | 3 rounds | 4 rounds | 5 rounds |
| 1. $A^P$ | 376 | 343 | 282 | 226.1 | 270.1 | 281.5 |
| 2a. $A^V$-EI | 409 | 387 | 364 | 226.3 | 304.5 | 350.2 |
| 2b. $A^V$-oSIA | 361 | 309 | 260 | 225.9 | 264.4 | 277.2 |
| 3. $A^M$ | 300 | 248 | 241 | 204.4 | 217.2 | 248.1 |

$A^P$ = Baseline allocation

**Figure 4:**
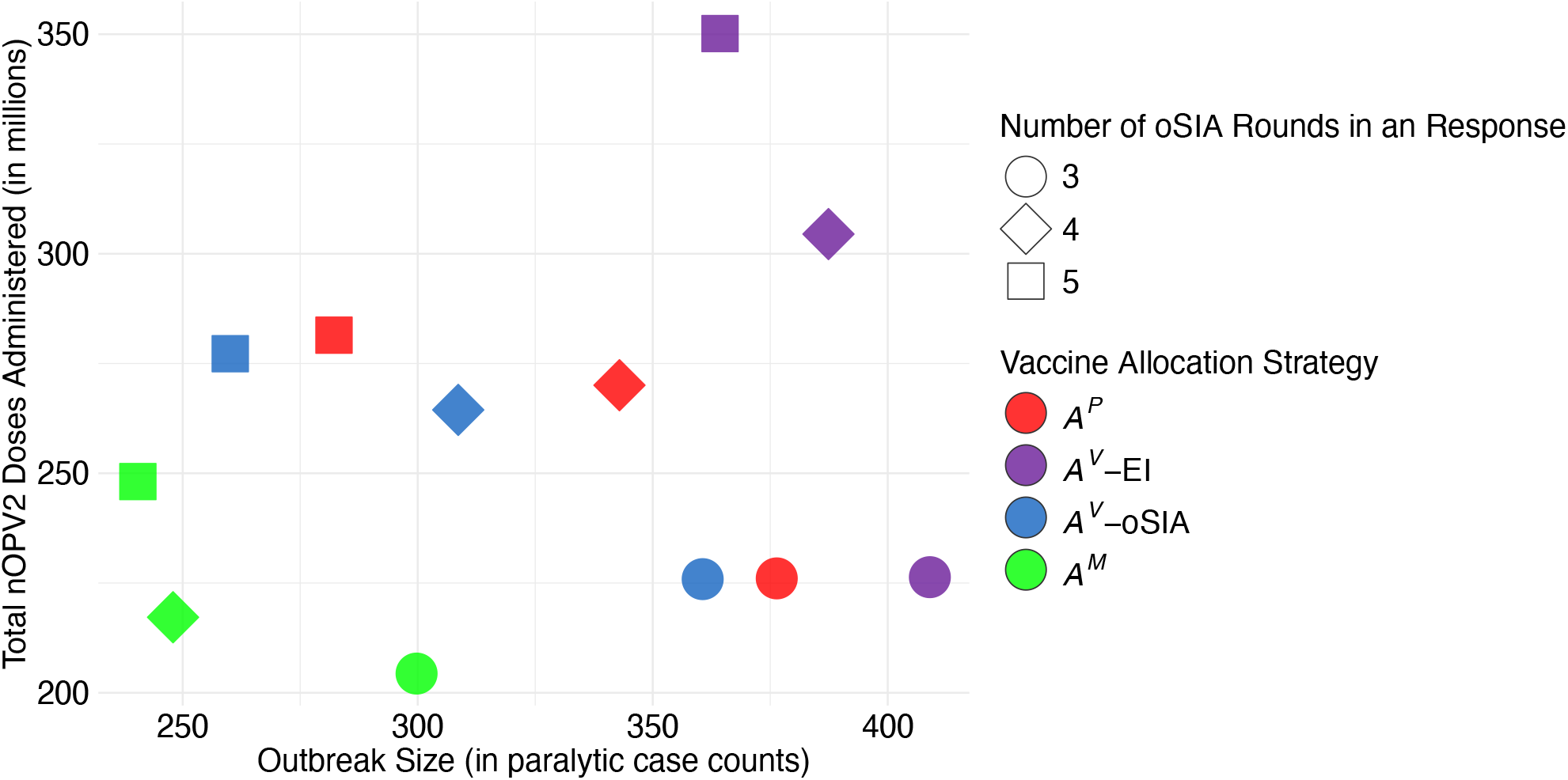
Comparison of outbreak size and total nOPV2 doses administered in tested outbreak response scenarios. *A*^*P*^= Baseline allocation; *A*^*V*^-EI = Priority allocation based on the history of IPV received through essential immunization; *A*^*V*^-oSIA = Priority allocation based on the history of OPV2 received through outbreak response supplementary immunization activities; *A*^*M*^ = Idealized allocation based on true immunity level

Compared with the baseline allocation of nOPV2 doses during oSIAs (*A*^*P*^), under a fixed number of oSIA rounds in each response, priority allocation based on the history of OPV2 received through oSIAs (*A*^*V*^-oSIA) resulted in smaller outbreak sizes with fewer nOPV2 doses administered in the response. In contrast, priority allocation based on the history of IPV received through essential immunization (*A*^*V*^-EI) resulted in larger outbreak sizes and higher number of nOPV2 doses used in the response. For example, with 4 oSIA rounds in each response, the *A*^*P*^strategy resulted in 343 paralytic cases using 270.1 million nOPV2 doses, whereas the *A*^*V*^-oSIA strategy resulted in 309 cases (a 10.0% reduction) using 264.4 million nOPV2 doses (a 2.1% reduction), and the *A*^*V*^-EI strategy resulted in 387 cases (a 13.0% increase) using 304.5 million nOPV2 doses (a 12.7% increase). Similar trend was observed for 3 and 5 rounds (see Table *3*)

For each vaccine allocation strategy, increasing the number of rounds in each response increased the number of nOPV2 doses used in oSIAs but reduced the outbreak size. For example, under the *A*^*V*^-EI strategy, the outbreak size decreased from 409 after 3 oSIA rounds to 364 after 5 rounds (11% reduction), with the number of doses used in oSIAs increased by 54.8%, from 226.3 to 350.2 million. Similar trend was observed for other vaccine allocation strategies.

## 4. Discussion

This study examined the extent to which improved operations of outbreak response could help completely interrupt cVDPV2 transmission. More specifically, we evaluated the impact of increasing the number of oSIA rounds implemented in each response and targeting under-vaccinated individuals during oSIAs based on individual’s vaccination history. To the best of our knowledge, this study stands as the first in polio modeling that (i) incorporates and simulates individual’s vaccination history, (ii) conceptualizes differences between low-immunity (based on true immunity achieved) and under-vaccinated (based on vaccine doses recorded) individuals, and (iii) offers unique insights into how the prioritization of under-vaccinated children influences the die-out dynamics of persistent poliovirus transmission.

Our simulations showed that cVDPV2 transmission was only eliminated in the most idealistic scenario, in which five oSIA rounds with ≥ 90% coverage were conducted in each response to the outbreaks and health workers could identify individuals with the lowest immunity levels and prioritize them for receiving nOPV2 doses during these rounds. In all other tested scenarios, which also included an ideal setting of oSIAs starting within 4 weeks of detection of transmission and reaching ≥ 90% coverage, cVDPV2 transmission persisted in all subpopulations at the end of the study period in 2028.

These findings underscore our most important insight: eliminating cVDPV2 transmission using nOPV2 only in outbreak response would require operational excellence that is far beyond what is being suggested by the current guidance and what has been achieved historically. The suggested number of oSIA rounds from our analysis is beyond the current guidelines which recommend two oSIA rounds that vaccinate children regardless of their vaccination histories [19]. The ideal timeliness and coverage that we assumed are beyond the performance of historical oSIAs since 2016, especially in Africa [8, 9, 24-26]. The prioritization of nOPV2 doses to those with low immunity levels, or even to those with zero or < 2 vaccine doses, is also extremely difficult to implement, especially in the settings with persistent cVDPV2 outbreaks. Not only we do not have methods to rapidly assess immunity levels before vaccination, but also vaccination cards documenting vaccines received through EI are often missing and it is non-existing for vaccines received through campaigns. Eliciting and recording information about previous vaccination would also limit the number of children that could be reached by a vaccination team per day and would increase the resources and time required to complete each vaccination round.

Our finding that increasing the number of oSIA rounds from three to five in each response led to smaller outbreak sizes, regardless of the vaccine allocation strategy, is significant. This aligns with previous modeling studies that emphasized the importance of additional vaccination rounds for accelerating outbreak control and ultimately stopping transmission [13, 16-18, 26-31]. Current GPEI guidelines recommend and fund two oSIA rounds in response to cVDPV2 outbreaks [19]. However, since 2016, only a small number of countries have succeeded in stopping outbreaks with just two rounds. This limited success is not surprising, as in many low-resource settings weak surveillance and outbreak response capacity delayed the detection of cVDPV2 transmission and made it difficult to mount timely campaigns with >90% coverage and adequately including under-vaccinated populations at highest risk of sustaining transmission.

We also demonstrated that using children’s OPV2 vaccination history from oSIAs to guide the nOPV2 allocation during outbreak response led to smaller outbreaks using fewer nOPV2 doses, whereas using IPV history from essential immunization did not provide the same benefit, compared with current guidelines that vaccinate all children regardless of prior vaccinations [19]. Further investigation showed that this difference arose because the effectiveness of using vaccination history depended on two factors (i) whether the history provided good alignment with the immunity against both infection and paralysis and (ii) whether low-immunity individuals are being prioritized while prioritizing under-vaccinated individuals (see Supplemental Materials S3). OPV2 records from oSIAs more accurately identified high-immunity children who acquired intestinal mucosal immunity. They marked those high-immunity children by high vaccination groups, which then made children in low vaccination groups were those truly with low immunity and were prioritized in subsequent oSIAs. In contrast, IPV records from essential immunization mainly reflected potential humoral immunity which poorly aligned with the immunity against infection and transmission. They tended to classify younger children into high vaccination groups, because they recently received IPV with high coverage, while older children, whose IPV doses were given years earlier with lower coverage, were classified into low vaccination groups. However, although IPV-vaccinated younger children were shielded from paralysis, they could still transmit poliovirus due to limited intestinal mucosal immunity while not being prioritized in oSIAs. On the other hand, older children, having experienced repeated exposures to cVDPV2 and nOPV2 campaigns, often had stronger intestinal mucosal immunity despite being labelled by low vaccination groups.

Prioritization of receiving nOPV2 during oSIAs based on true immunity consistently outperformed vaccine allocation strategies based on vaccination history, which underlines the limitations of history of vaccines received through essential immunization or outbreak response to serve as proxies for the immunity against both infection and paralysis. Vaccination history reflects only potential protection achieved by administered doses. The identification of fully susceptible and low-immunity individuals for vaccine prioritization would require not only accurate information on vaccine type, number of doses, and time since vaccination, but also knowledge of historical transmission and vaccine effectiveness in the area, which remains difficult in real life.

Altogether, our modeling results indicate that in the absence of highly sophisticated and extremely robust outbreak response, eliminating cVDPV2 through outbreak response alone remains extremely challenging. Similar to the findings from other studies [27, 32-35], our results support incorporating OPV2 into various preventive vaccination programs, such as essential immunization, preventive SIAs, and continuous vaccination, to enhance the control of cVDPV2 transmission. In essential immunization, nOPV2 could be added to the existing schedule of bOPV and IPV, either through concomitant administration or separating bOPV and nOPV2 [36]. Preventive SIAs may use nOPV2 to vaccinate children across a broad age range, thereby enhancing population immunity against type 2 poliovirus and preventing its transmission. Continuous vaccination strategies, such as administering nOPV2 during health facility visits or at border crossings in high-risk regions, can effectively reach under-vaccinated children in conflict-affected areas and highly mobile populations, as demonstrated by the “Permanent Transit Point for Polio” strategy at the Pakistan–Afghanistan border [37].

Our study has several limitations. First, we adapted a deterministic compartmental model of poliovirus transmission [16, 17], which has its limitations. These include the lack of stochastic elements – such as random variation in infectious contacts – that can influence the interruption of transmission. The model uses large subpopulations, which may overlook important real-world heterogeneities, such as changes in mixing patterns within subpopulations, that can significantly affect community-level virus transmission. Also, the model assumes constant accessibility to all tested subpopulations in Nigeria. Future work should consider dynamic accessibility given data availability. Second, the model does not consider vaccine dose over-reporting. However, reports of fake finger marks in countries such as Nigeria [unpublished data; personal communication with Oladayo Biya] and Pakistan [38] have indicated that some individuals’ vaccination histories may be overestimated. Over-reporting can misclassify low-immunity individuals as well-vaccinated, preventing them from being prioritized in outbreak response that target the under-vaccinated. Future work can examine how over-reporting affects the outcomes of priority allocation based on recorded vaccination history. Third, the model does not incorporate logistical and operational constraints that may affect the feasibility of vaccine allocation strategies prioritizing under-vaccinated children. When individual-level data are not available to identify these children, alternative approaches can be explored to guide resource allocation toward under-vaccinated communities, such as those in remote or conflict areas, highly mobile groups, or socially marginalized populations. Practical strategies to support this include the geographic information system tracking of vaccinators [39]. Even in an idealistic situation where individual-level vaccination records are accessible, further studies are needed to assess the costs and cost-effectiveness of such strategies prioritizing under-vaccinated children given limited financial resources [40]. Together, these conditions underscore that our analysis is not definitive; rather, it should be viewed as a step toward additional work needed to evaluate and improve vaccination strategies under a range of real-world conditions. Finally, our model tracks and records only vaccine doses given to susceptible individuals for simplicity, as doses given to exposed, infectious, or IPV-reacted individuals have no effects in the model. In reality, vaccine doses will be administered also on non-susceptible individuals, who are often asymptomatic and therefore hard to distinguish from those who are susceptible. To account for this, one could track all vaccine doses, regardless of recipients’ susceptibility. However, while this approach will increase the total number of nOPV2 doses administered, it will not alter our main findings on how outbreak outcomes are affected by the number of oSIA rounds and the vaccine allocation strategy.

## 5. Conclusions

Our study shows that improving the operations of outbreak response, by adding more oSIA rounds and prioritizing under-vaccinated children based on the history of OPV2 doses received through campaigns, can reduce cVDPV2 paralytic case counts while using fewer nOPV2 doses compared with the current strategy outlined in the standard operating procedures [19]. However, these improvements alone are not sufficient for complete interruption of cVDPV2 transmission in high-risk settings such as Nigeria. Achieving interruption would require exceptionally high-level operational performance: at least five timely (within 4 weeks) and high coverage (90%) nOPV2 oSIA rounds for each outbreak response, all capable of accurately identifying and targeting truly low-immunity children. This level of performance remains far beyond what has historically been feasible and represents a highly sophisticated and idealized scenario.

These findings highlight an important consideration for stakeholders: strengthening outbreak response alone through an improvement in the number of rounds, vaccine allocation strategies, and timeliness and coverage of campaigns may not be sufficient or practical to completely interrupt persistent cVDPV2 outbreaks. A more practical and sustainable path forward may involve combining reactive outbreak response vaccinations with preventive vaccinations such as the inclusion of nOPV2 in essential immunization. Pairing the preventive and reactive uses of nOPV2 may provide a more promising approach leading to the elimination of cVDPV2 and the achievement of global eradication goals.

## Data Availability

All of the data that the authors can share is available in the public domain and appropriate citations are provided.

## Declarations

### Financial support

This study was funded by a cooperative agreement between the Georgia Institute of Technology Tech and the Centers for Diseases Control and Prevention (1 NU2RGH001919-01-00). This research was also supported in part by the Center for Health and Humanitarian Systems, William W. George endowment, Harold R. and Mary Anne Nash endowment, and George Family Fellowship at Georgia Tech, and the INFORMS Seth Bonder Scholarship.

### Conflicts of interest

The authors declare that they have no known conflicts of interest or personal relationships that could have appeared to influence the work in this paper.

### Consent for publication

All authors have approved the final article for publication.

### Code availability

Upon request

## Acknowledgements

We thank the Nigeria National Primary Health Care Development Agency for sharing the polio data. We appreciate the funding support to this study from the Centers for Disease Control and Prevention (1 NU2RGH001919-01-00), the Center for Health and Humanitarian Systems, William W. George endowment, Harold R. and Mary Anne Nash endowment, and George Family Fellowship at the Georgia Institute of Technology, and the INFORMS Seth Bonder Scholarship. The findings and conclusions in this report are those of the authors and do not necessarily represent the views of the Centers for Disease Control and Prevention. All authors attest they meet the ICMJE criteria for authorship.

## Notes

### Competing Interest Statement

The authors declare the following financial interests/personal relationships which may be considered as potential competing interests: Yuming Sun reports financial support was provided by Centers for Disease Control and Prevention. Pinar Keskinocak reports financial support was provided by Centers for Disease Control and Prevention. Lauren N. Steimle reports financial support was provided by Centers for Disease Control and Prevention. Stephanie D. Kovacs reports a relationship with Centers for Disease Control and Prevention that includes: employment.

### Summary of Updates

Revise the typo in Title. It should be "Quantifying" instead of "Qualifying."

