## Supplemental Materials for "Quantifying the benefits of improved operations for polio outbreak response: A model-based analysis"

### S1. Updated general live poliovirus transmission model

In this section, we show the updates added to the existing model and the necessary details of the model to better understand our study (e.g., feasible transitions from immunity group $i$ to immunity group $i'$ in Table S1). Additional details of the model can be found in [1, 2]. Of notice, the model is general and can be applied to simulate the transmission of any serotypes in any populations. It is not limited by the current vaccination programs in Nigeria that deliver nOPV2 only through outbreak response and have no OPV2 in essential immunization.

#### S1.1. Updated notations

$i$ = immunity group ($i = 0,1,\ldots,7$)

- $i = 0$: unimmunized
- $i = 1$: IPV-only-immunized; received the most recent effective IPV dose more than two years ago; $\geq 1$ effective IPV dose and no live poliovirus exposures in life
- $i = 2$: IPV-only-immunized; received the most recent effective IPV dose within the past two years; only 1 effective IPV dose and no live poliovirus exposures in life
- $i = 3$: IPV-only-immunized; received the most recent effective IPV dose within the past two years; only 2 effective IPV doses and no live poliovirus exposures in life
- $i = 4$: IPV-only-immunized; received the most recent effective IPV dose within the past two years; $\geq 3$ effective IPV doses and no live poliovirus exposures in life
- $i = 5$: Live-poliovirus-immunized; most recently acquired immunity more than two years ago; $\geq 1$ live poliovirus exposure with or without effective IPV doses in life
- $i = 6$: Live-poliovirus-immunized; most recently acquired immunity within the past two years; only 1 live poliovirus exposure and no effective IPV doses in life
- $i = 7$: Live-poliovirus-immunized; most recently acquired immunity within the past two years; $\geq 2$ live poliovirus exposures without effective IPV doses in life or both live poliovirus exposure(s) and effective IPV dose(s) in life

$k$ = vaccination group ($k = 0,1,\ldots,7$)

- $k = 0$: unvaccinated
- $k = 1$: IPV-only-vaccinated; received the most recent IPV dose more than two years ago; $\geq 1$ IPV dose and no OPV doses in life
- $k = 2$: IPV-only-vaccinated; received the most recent IPV dose within the past two years; only 1 IPV dose and no OPV doses in life
- $k = 3$: IPV-only-vaccinated; received the most recent IPV dose within the past two years; only 2 IPV doses and no OPV doses in life
- $k = 4$: IPV-only-vaccinated; received the most recent IPV dose within the past two years; $\geq 3$ IPV doses and no OPV doses in life
- $k = 5$: OPV-vaccinated; received the most recent poliovirus vaccine dose more than two years ago; $\geq 1$ OPV dose with or without IPV doses in life
- $k = 6$: OPV-vaccinated; received the most recent poliovirus vaccine dose within the past two years; only 1 OPV dose with or without IPV doses in life

- $k = 7$: OPV-vaccinated; received the most recent poliovirus vaccine doses within the past two years; $\geq 2$ OPV doses without IPV doses in life or both OPV and IPV doses in life

$j$ = virus strain ($j = 1, \ldots, 20$)
- $j = 1$: original virus in nOPV or Sabin-strain OPV (e.g., trivalent OPV, bivalent OPV, and monovalent OPV)
- $j = 2 - 19$: partially reverted forms of the original virus in Sabin-strain OPV or nOPV
- $j = 20$: fully reverted form of the original virus, i.e., cVDPV

$S_{i,k,a,s}(t)$ = susceptible individuals in immunity group $i$, vaccination group $k$, age group $a$, and subpopulation $s$ at time $t$

$H_{i,k,a,s}(t)$ = IPV-reacted individuals in immunity group $i$, vaccination group $k$, age group $a$, and subpopulation $s$ at time $t$

$E_{i,k,j,a,s}(t)$ = exposed individuals infected by virus strain $j$ in immunity group $i$, vaccination group $k$, age group $a$, and subpopulation $s$ at time $t$

$I_{i,k,j,a,s}(t)$ = infectious individuals infected by virus strain $j$ in immunity group $i$, vaccination group $k$, age group $a$, and subpopulation $s$ at time $t$

$tr_{a,s}^{R,v}$ = per-dose effectiveness of vaccine $v$ in the essential immunization of age group $a$ and subpopulation $s$ (vaccine $v = 0$ (nOPV), 1 (Sabin-strain OPV), 2 (IPV))

$cov_{a,s}^{R,v}(t)$ = coverage of vaccine $v$ in the essential immunization of age group $a$ and subpopulation $s$ at time $t$

$e_{a,s}^{v}(t)$ = effective vaccination percentage of vaccine $v$ in the essential immunization of age group $a$ and subpopulation $s$ at time $t$

$u_{a,s}^{v}(t)$ = ineffective vaccination percentage of vaccine $v$ in the essential immunization of age group $a$ and subpopulation $s$ at time $t$ (vaccine $v = 0$ (nOPV), 1 (Sabin-strain OPV), 2 (IPV))

$\zeta_{a,s}^{v}$ = probability of recording vaccine $v$ of essential immunization in age group $a$ and subpopulation $s$ for individual's vaccination history (in this study, we considered only the values 0 and 1)

$\tau_{s}^{st,S,v,r}$ = start day of the $r$-th round of vaccine $v$ SIA in subpopulation $s$

$\tau_{s}^{en,S,v,r}$ = end day of the $r$-th round of vaccine $v$ SIA in subpopulation $s$

$d_{s}^{v,r}$ = duration of the $r$-th round of vaccine $v$ SIA in subpopulation $s$

$tag_{s}^{v,r}$ = target age groups of the $r$-th round of vaccine $v$ SIA in subpopulation $s$

$cov_{a,s}^{S,v,r}$ = coverage of the $r$-th round of vaccine $v$ SIA in subpopulation $s$

$tr_{s}^{S,v,r}$ = per-dose effectiveness of the vaccine used in the $r$-th round of vaccine $v$ SIA in subpopulation $s$

$f_{a,s}^{v}(t)$ = effective coverage of vaccine $v$ through SIAs in age group $a$ and subpopulation $s$ at time $t$

$o_{a,s}^{v}(t)$ = ineffective coverage of vaccine $v$ through SIAs in age group $a$ and subpopulation $s$ at time $t$

$\iota_{a,s}^{v}$ = probability of recording vaccine $v$ of SIAs in age group $a$ and subpopulation $s$ for individual's vaccination history (in this case study, we considered only the values 0 and 1)

#### S1.2. Updated equations

$$e_{a,s}^{v}(t) = cov_{a,s}^{R,v}(t) \times tr_{a,s}^{R,v}$$

$$u_{a,s}^{v}(t) = cov_{a,s}^{R,v}(t) \times \left(1 - tr_{a,s}^{R,v}\right)$$

$$f_{a,s}^{v}(t) = 1_{\left\{\tau_s^{st,S,v,r} \le t \le \tau_s^{en,S,v,r},\ a \in tag_s^{v,r}\right\}} \times \frac{-\log\left(1 - cov_{a,s}^{S,v,r} \times tr_s^{S,v,r}\right)}{d_s^{v,r}}$$

$$o_{a,s}^{v}(t) = 1_{\left\{\tau_s^{st,S,v,r} \le t \le \tau_s^{en,S,v,r},\ a \in tag_s^{v,r}\right\}} \times \left[\frac{-\log\left(1 - cov_{a,s}^{S,v,r}\right)}{d_s^{v,r}} - \frac{-\log\left(1 - cov_{a,s}^{S,v,r} \times tr_s^{S,v,r}\right)}{d_s^{v,r}}\right]$$

#### S1.3. Updated OPV reversion

We referred to the live-attenuated virus in Sabin-strain OPV and nOPV as Sabin-strain OPV virus and nOPV virus, respectively.

Reversion of the Sabin-strain OPV virus follows the same as in previous studies [1-3]. 20 progressive reversion stages (i.e., virus strains) are considered for the Sabin-strain OPV virus, from stage 1 for the Sabin-strain OPV virus, to stages 2-19 for the partially reverted forms, and to stage 20 for the fully reverted form (e.g., cVDPV). These 20 reversion stages are differentiated in the model by the transmissibility (i.e., basic reproductive number $R_0$) and the neurovirulence (i.e., paralysis to infection rate $PIR$) of the virus strains they represent. Assuming that $R_{0_j}^{avg}$ is the average $R_0$ of virus strain $j$ in a calendar year, $R_{0_1}^{avg}$ and $R_{0_{20}}^{avg}$ are given for the Sabin-strain OPV virus and the fully reverted form, respectively, the model calculates $R_{0_j}^{avg}$ for each virus strain $j$ based on $R_{0_j}^{avg} = R_{0_{20}}^{avg} - \left(R_{0_{20}}^{avg} - R_{0_1}^{avg}\right) \times \left(\frac{20-j}{19}\right)$ [4]. As reversion is a process where the Sabin-strain OPV virus regains transmissibility and neurovirulence, $R_{0_j}^{avg}$ increases when $j$ increases in the model, for $j = 1,2,\ldots,20$.

The model assumes that the nOPV virus has the same transmissibility as the Sabin-strain OPV virus of the same serotype in the studied population, i.e., $R_{0_1}^{avg}$. Moreover, as the model assumes that the nOPV virus reverts but at a reduced level compared to the Sabin-strain OPV virus [5], the first 10 reversion stages (i.e., virus strains 1-10) of nOPV remain the same as reversion stage 1, and the last 10 reversion stages of nOPV virus are the same as the last 10 reversion stages for Sabin-strain OPV virus reversion. Figure S1 provides an illustration on the value of $R_{0_j}^{avg}$ of each reversion stage (or virus strain) of Sabin-strain OPV and nOPV, in the case study of cVDPV2 transmission in Nigeria.

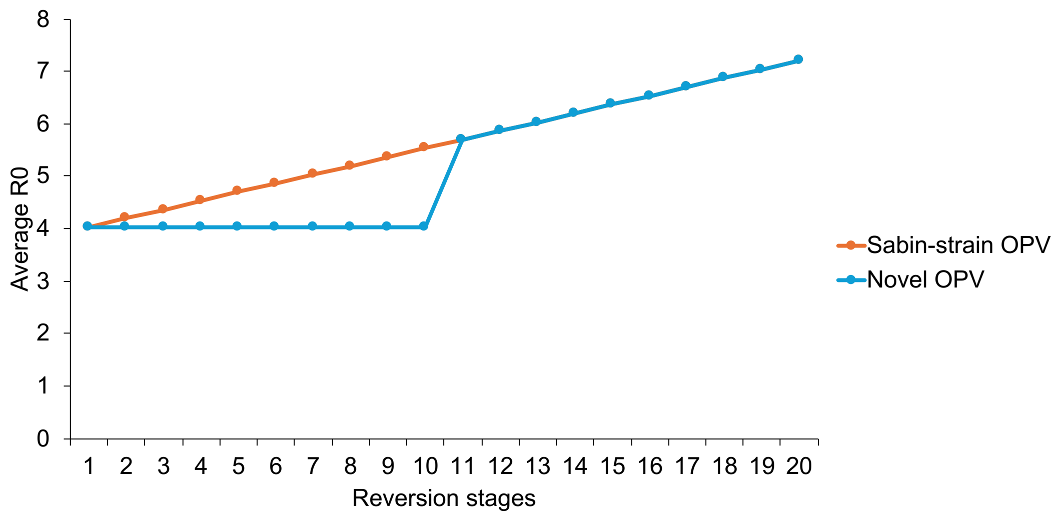


Figure S1: Average $R_0$ values of different reversion stages of Sabin-strain oral poliovirus vaccine (OPV) and novel OPV

The model sets up a positive paralysis to infection rate (e.g., 1 over 2000 for serotype 2 in the case study of Nigeria) for virus strain 20 and assigns zeros for virus strains 1-19, as it considers no vaccine-associated paralytic poliomyelitis (VAPP).

#### S1.4. Updated transitions caused by changes of vaccination groups

Due to the addition of vaccination groups to the live poliovirus transmission model, possible transitions are updated to capture changes in vaccination groups caused by different vaccination programs and timing of the most recent vaccination.

**Vaccination through essential immunization:** Essential immunization is a regular and preventative vaccination program that vaccinates individuals when they reach certain ages. In the model, individuals of $E_{i,k,j,a-1,s}$, $I_{i,k,j,a-1,s}$, and $H_{i,k,a-1,s}$ transition to $E_{i,k,j,a,s}$, $I_{i,k,j,a,s}$, and $H_{i,k,a,s}$, respectively, with the rate $1/w_{a-1}$ (for $a = 2, \dots, n_a$) where $w_a$ is defined as the number of days contained by age group $a$ (e.g., $w_{a=1} = 365 * 5 = 1825$ days if $a = 1$ is for children aged 0-4 years).

Denote $e_{a,s}^{v}$ and $u_{a,s}^{v}$ as the effective and ineffective vaccination percentage of essential immunization in subpopulation $s$ that uses vaccine $v$ ($v =$ 0-2 where 0 represents nOPV, 1 represent Sabin-strain OPV, and 2 represents IPV) on susceptible individuals when they reach age group $a$, respectively. Both $e_{a,s}^{v}$ and $u_{a,s}^{v}$ depend on coverage and effectiveness of vaccine $v$ in essential immunization (see equations in Supplemental Materials S1.2). Denote $\zeta_{a,s}^{v}$ as the record probability of vaccine $v$ in essential immunization for subpopulation $s$ and age group $a$. Among individuals in $S_{i,k,a-1,s}$ that transition to age group $a$ and receive effective vaccine doses, proportions $e_{a,s}^{0} \times \zeta_{a,s}^{0}$, $e_{a,s}^{1} \times \zeta_{a,s}^{1}$, and $e_{a,s}^{2} \times \zeta_{a,s}^{2}$ of them transition to $E_{i,k',21,a,s}$, $E_{i,k',1,a,s}$, and $H_{i,k',a,s}$ (see Table S2 for feasible transitions from vaccination group $k$ to vaccination group $k'$), respectively, due to recording vaccine doses, and proportions $e_{a,s}^{0} \times (1 - \zeta_{a,s}^{0})$, $e_{a,s}^{1} \times (1 - \zeta_{a,s}^{1})$, and $e_{a,s}^{2} \times (1 - \zeta_{a,s}^{2})$ of them transition to $E_{i,k,21,a,s}$, $E_{i,k,1,a,s}$, and $H_{i,k,a,s}$, respectively, due to not recording vaccine doses. Among individuals in $S_{i,k,a-1,s}$ that transition to age group $a$ and receive ineffective vaccine doses, proportions $u_{a,s}^0 \times \zeta_{a,s}^0$, $u_{a,s}^1 \times \zeta_{a,s}^1$, and $u_{a,s}^2 \times \zeta_{a,s}^2$ of them transition to $S_{i,k',a+1,s}$ due to recording vaccine doses where $k'$ can be different given vaccine $v$. The rest of those individuals in $S_{i,k,a-1,s}$ that transition to age group $a$ transition to $S_{i,k,a,s}$, i.e., those who receive no vaccines in essential immunization or receive ineffective doses that are not recorded.

Similarly, among newborns of subpopulation $s$ that enter the model based on the birth rate $b$: proportions $e_{1,s}^0 \times \zeta_{1,s}^0$, $e_{1,s}^1 \times \zeta_{1,s}^1$, and $e_{1,s}^2 \times \zeta_{1,s}^2$ of them transition to $E_{0,6,21,1,s}$, $E_{0,6,1,1,s,}$, and $H_{0,2,1,s}$, respectively, due to effective doses with recording; proportions $e_{1,s}^0 \times (1 - \zeta_{1,s}^0)$, $e_{1,s}^1 \times (1 - \zeta_{1,s}^1)$, and $e_{1,s}^2 \times (1 - \zeta_{1,s}^2)$ of them transition to $E_{0,0,21,1,s}$, $E_{0,0,1,1,s}$, and $H_{0,0,1,s}$, respectively, due to effective doses with no recording; proportions $u_{1,s}^0 \times \zeta_{1,s}^0$, $u_{1,s}^1 \times \zeta_{1,s}^1$, and $u_{1,s}^2 \times \zeta_{1,s}^2$ of them transition to $S_{0,6,1,s}$, $S_{0,6,1,s}$, and $S_{0,2,1,s}$, respectively, due to ineffective doses with recording; and the rest of them transition to $S_{0,0,1,s}$ due to no vaccines received or receiving ineffective doses with no recording.

Table S1: Feasible transitions from immunity group $i$ to immunity group $i'$

| | $i' = 0$ | $i' = 1$ | $i' = 2$ | $i' = 3$ | $i' = 4$ | $i' = 5$ | $i' = 6$ | $i' = 7$ |
|---|---|---|---|---|---|---|---|---|
| $i = 0$ | | | $H$ to $S$ | | | | $I$ to $S$ | |
| $i = 1$ | | | | $H$ to $S$ | | | | $I$ to $S$ |
| $i = 2$ | | $S$ to $S$ | | $H$ to $S$ | | | | $I$ to $S$ |
| $i = 3$ | | $S$ to $S$ | | | $H$ to $S$ | | | $I$ to $S$ |
| $i = 4$ | | $S$ to $S$ | | | $H$ to $S$ | | | $I$ to $S$ |
| $i = 5$ | | | | | | | | $I/H$ to $S$ |
| $i = 6$ | | | | | | $S$ to $S$ | | $I/H$ to $S$ |
| $i = 7$ | | | | | | $S$ to $S$ | | $I/H$ to $S$ |

**Vaccination through supplementary immunization activities (SIAs):** SIAs are short-term (e.g., 4 days) and periodic vaccination programs that vaccinate a large age group of children (e.g., 0-4 years) to prevent (i.e., preventive SIAs or pSIAs) or respond to (i.e., outbreak response SIAs or oSIAs) outbreaks. In the model, we denote $f_{a,s}^v$ and $o_{a,s}^v$ as the effective and ineffective coverage of vaccine $v$ through SIAs in age group $a$ and subpopulation $s$, respectively. Both $f_{a,s}^v$ and $o_{a,s}^v$ depend on implementation period and coverage of SIAs using vaccine $v$ in age group $a$ and subpopulation $s$, and the effectiveness of vaccine $v$ (see equations in Supplemental Materials S1.2). Denote $\iota_{a,s}^v$ as the probability of recording vaccine $v$ of SIAs in age group $a$ and subpopulation $s$ for individuals' recorded vaccination histories ($v = 0,1,2$). Therefore, given effective doses with recording, individuals in $S_{i,k,a,s}$ transition to $E_{i,k',21,a,s}$ , $E_{i,k',1,a,s}$, and $H_{i,k',a,s}$, based on $f^0_{a,s} \times \iota^0_{a,s}$, $f^1_{a,s} \times \iota^1_{a,s}$, and $f^2_{a,s} \times \iota^2_{a,s}$, respectively. Given effective doses with no recording, individuals in $S_{i,k,a,s}$ transition to $E_{i,k,21,a,s}$ , $E_{i,k,1,a,s}$, and $H_{i,k,a,s}$, based on $f^0_{a,s} \times (1 - \iota^0_{a,s})$, $f^1_{a,s} \times (1 - \iota^1_{a,s})$, and $f^2_{a,s} \times (1 - \iota^2_{a,s})$, respectively. Given ineffective doses with recording, individuals in $S_{i,k,a,s}$ transition to $S_{i,k',a,s}$ based on $o^v_{a,s} \times \iota^v_{a,s}$ where $k'$ can be different given $k$ and $v$.

**Timing of the most recent vaccination:** Since the characterizations of vaccination groups separates vaccines received within the past two years and at least two years ago, the model captures this dynamic by having individuals in $S_{i,k,a,s}$ transition to $S_{i,k'a,s}$ after two years. This is similar to the waning of immunity in [1, 2] but the "waning" of vaccination history.

Table S2: Feasible transitions from vaccination group $k$ to vaccination group $k'$

| | $k' = 0$ | $k' = 1$ | $k' = 2$ | $k' = 3$ | $k' = 4$ | $k' = 5$ | $k' = 6$ | $k' = 7$ |
|---|---|---|---|---|---|---|---|---|
| $k = 0$ | | | $S$ to $S/H$ | | | | $S$ to $S/E$ | |
| $k = 1$ | | | | $S$ to $S/H$ | | | | $S$ to $S/E$ |
| $k = 2$ | | $S$ to $S$ | | $S$ to $S/H$ | | | | $S$ to $S/E$ |
| $k = 3$ | | $S$ to $S$ | | | $S$ to $S/H$ | | | $S$ to $S/E$ |
| $k = 4$ | | $S$ to $S$ | | | $S$ to $S/H$ | | | $S$ to $S/E$ |
| $k = 5$ | | | | | | | | $S$ to $S/E/H$ |
| $k = 6$ | | | | | | $S$ to $S$ | | $S$ to $S/E/H$ |
| $k = 7$ | | | | | | $S$ to $S$ | | $S$ to $S/E/H$ |

### S2. Initial condition

We provide in this section additional details on the estimation of initial condition for Northwest and Northeast Nigerian population's distribution across vaccination groups at the start of 2024.

Table S3 presents the estimates on the coverages of receiving at least one IPV dose (i.e., IPV1) and for receiving at least two IPV doses (i.e., IPV2) in children aged 0-4 years at the start of 2024 in all five non-isolated subpopulations.

The Institute for Health Metrics and Evaluation (IHME) provides estimates on DTP1 (i.e., coverage for receiving at least one Diphtheria, Tetanus, and Pertussis (DTP) vaccine dose) and DTP3 (coverage for receiving at least three DTP vaccine doses) of each birth cohort of 0-4 years in each state of Northwest and Northeast Nigeria. Nigeria's Demographic and Health Survey (DHS) 2024 provides IPV1 and IPV2 estimates for the 2023 birth cohort (i.e., children aged $< 1$ year in the initial condition) in each state of Northwest and Northeast Nigeria.

Given that first (second) IPV dose is given together with first (third) DTP dose, we estimated IPV1 (IPV2) for children aged 1, 2, 3, and 4 years in each state of Northwest and Northeast Nigeria, based on (i) proportion of IPV1 (IPV2) to DTP1 (DTP3) of children aged $< 1$ year (the 2023 birth cohort) and (ii) DTP1 (DTP3) of children aged 1, 2, 3, and 4 years. For example, in Jigawa, DTP1 and IPV1 for children aged $< 1$ year are 67% and 62.3%, respectively. As DTP1 was estimated as 66.2% by IHME for children aged 1 year in Jigawa, we estimated IPV1 for this birth cohort to be $(\frac{62.3\%}{67\%}) \times 66.2\% = 61.5\%$. Since the second IPV dose was introduced in Nigeria in July 2021 [6], we reduced IPV2 estimates for children aged 2 years (the 2021 birth cohort) by 60% and that of children aged 3 and 4 years (the 2020 and 2019 birth cohorts, respectively) to 0. State-level estimates of IPV1 and IPV2 were then aggregated into a subpopulation by taking the weighted average of estimates of all states included in that subpopulation (see [1, 2] for more details).

Using these IPV1 and IPV2 estimates, we estimated the proportions of children aged 0-4 years in vaccination group 0 (unvaccinated) and vaccination groups 1-3 (IPV-only-vaccinated but no more than 2 IPV doses), as shown in Table S4.

As we assumed that essential immunization happens when children reached the age of 3 months in the model for Nigeria, all children aged 0-2 months were characterized by vaccination group 0, i.e., unvaccinated. For children aged 3-11 months, the proportions of individuals in vaccination group 3 (IPV only; received the most recent IPV dose within the past 2 years and had 2 IPV doses in life), vaccination group 2 (IPV only; received the most recent IPV dose within the past 2 years and had 1 IPV dose in life), and vaccination group 0 are IPV2, IPV1−IPV2, and $100\% -$ (IPV1+IPV2), respectively. The proportions of individuals in all other vaccination groups are 0.

For children aged 2-4 years, none of them should be in vaccination groups representing those who received the most recent IPV dose within the past 2 years, and therefore we set up zero proportions for vaccination groups 2-4. The proportions for vaccination groups 0 and 1 are IPV1 and $100\%$ −IPV1, respectively. The proportions of individuals in all other vaccination groups are 0.

We didn't estimate IPV1 and IPV2 for individuals aged $\geq$ 5 years. We assumed a simplest case here where all of them were in vaccination group 0. The distribution of children aged $\geq$ 5 years does not impact the outcomes of vaccine allocation strategies, as the target ages of all oSIAs are 0-4 years.

Also, we did not estimate the history of OPV2 doses received through outbreak response in the initial condition, as no data is available to gauge the individual-level vaccination status of OPV2. We assumed all simulations started with no individuals in OPV-vaccinated groups 5-7.

Table S3: Estimated IPV1 (coverage for receiving at least one inactivated poliovirus vaccine (IPV) dose) and IPV2 (coverage for receiving at least two IPV doses) for children aged 0-4 years in subpopulations 1-5

| Age groups | Birth cohorts | Subpopulation 1 | | Subpopulation 2 | | Subpopulation 3 | | Subpopulation 4 | | Subpopulation 5 | |
|---|---|---|---|---|---|---|---|---|---|---|---|
| | | IPV1* | IPV2* | IPV1 | IPV2 | IPV1 | IPV2 | IPV1 | IPV2 | IPV1 | IPV2 |
| 0 years | 2023 | 51.8% | 36.2% | 34.8% | 21.3% | 58.5% | 40.8% | 55.1% | 41.6% | 37.3% | 22.1% |
| 1 year | 2022 | 50.9% | 35.2% | 34.5% | 20.8% | 58.2% | 40.3% | 55.1% | 41.4% | 38.6% | 22.9% |
| 2 years | 2021 | 50.1% | 13.5% | 36.7% | 8.3% | 58.2% | 15.8% | 51.2% | 14.9% | 37.2% | 8.8% |
| 3 years | 2020 | 48.3% | 0.0% | 38.1% | 0.0% | 57.0% | 0.0% | 47.5% | 0.0% | 34.9% | 0.0% |
| 4 years | 2019 | 45.8% | 0.0% | 37.5% | 0.0% | 54.7% | 0.0% | 45.6% | 0.0% | 34.8% | 0.0% |

*IPV1 and IPV2 are estimated based on estimated IPV coverage from Nigeria's Demographic and Health Survey 2023-2024 [7] and estimated DTP coverage from the Institute for Health Metrics and Evaluation [8]

Table S4: Estimated proportions of children aged 0-4 years (at the start of 2024) in vaccination groups 0-3 for subpopulations 1-5

| Age groups | Birth cohorts | Vaccination groups* | | | |
|---|---|---|---|---|---|
| | | 0 | 1 | 2 | 3 |
| | | ***Subpopulation 1*** | | | |
| 0-2 months | 2023 | 100.0% | 0.0% | 0.0% | 0.0% |
| 3-11 months | 2023 | 48.2% | 0.0% | 15.7% | 36.2% |
| 1 year | 2022 | 49.1% | 0.0% | 15.7% | 35.2% |
| 2 years | 2021 | 49.9% | 50.1% | 0.0% | 0.0% |
| 3 years | 2020 | 51.7% | 48.3% | 0.0% | 0.0% |
| 4 years | 2019 | 54.2% | 45.8% | 0.0% | 0.0% |
| | | ***Subpopulation 2*** | | | |
| 0-2 months | 2023 | 100.0% | 0.0% | 0.0% | 0.0% |
| 3-11 months | 2023 | 65.2% | 0.0% | 13.5% | 21.3% |
| 1 year | 2022 | 65.5% | 0.0% | 13.8% | 20.8% |
| 2 years | 2021 | 63.3% | 36.7% | 0.0% | 0.0% |
| 3 years | 2020 | 61.9% | 38.1% | 0.0% | 0.0% |

| | | | | | |
|---|---|---|---|---|---|
| 4 years | 2019 | 62.5% | 37.5% | 0.0% | 0.0% |
| | | ***Subpopulation 3*** | | | |
| 0-2 months | 2023 | 100.0% | 0.0% | 0.0% | 0.0% |
| 3-11 months | 2023 | 41.5% | 0.0% | 17.7% | 40.8% |
| 1 year | 2022 | 41.8% | 0.0% | 17.9% | 40.3% |
| 2 years | 2021 | 41.8% | 58.2% | 0.0% | 0.0% |
| 3 years | 2020 | 43.0% | 57.0% | 0.0% | 0.0% |
| 4 years | 2019 | 45.3% | 54.7% | 0.0% | 0.0% |
| | | ***Subpopulation 4*** | | | |
| 0-2 months | 2023 | 100.0% | 0.0% | 0.0% | 0.0% |
| 3-11 months | 2023 | 44.9% | 0.0% | 13.5% | 41.6% |
| 1 year | 2022 | 44.9% | 0.0% | 13.7% | 41.4% |
| 2 years | 2021 | 48.8% | 51.2% | 0.0% | 0.0% |
| 3 years | 2020 | 52.5% | 47.5% | 0.0% | 0.0% |
| 4 years | 2019 | 54.4% | 45.6% | 0.0% | 0.0% |
| | | ***Subpopulation 5*** | | | |
| 0-2 months | 2023 | 100.0% | 0.0% | 0.0% | 0.0% |
| 3-11 months | 2023 | 62.7% | 0.0% | 15.2% | 22.1% |
| 1 year | 2022 | 61.4% | 0.0% | 15.7% | 22.9% |
| 2 years | 2021 | 62.8% | 37.2% | 0.0% | 0.0% |
| 3 years | 2020 | 65.1% | 34.9% | 0.0% | 0.0% |
| 4 years | 2019 | 65.2% | 34.8% | 0.0% | 0.0% |

*Proportions in vaccination groups 0-3 are estimated based on IPV1 and IPV2 estimated in Table S3

### S3. Additional results

To compare how essential immunization and oSIA vaccination histories impacted the changes in vaccination groups and provided different inferences into true immunity level, we measured the "mapping between immunity and vaccination groups." The mapping between immunity and vaccination groups in subpopulation $s$ at time $t$ shows the percentage of subpopulation $s$' individuals aged 0-4 years in each combination of an immunity group and a vaccination group at time $t$.

Figure S2 shows the mapping between immunity and vaccination groups in subpopulations 1-5, at the start days of simulations (Day 1) and of the 5 subsequent oSIA rounds (SIAs 1-5) in the first outbreak response, under strategies $A^V$-EI and $A^V$-oSIA. Each big square represents the mapping on a specific day. Each small square in a big square represents a combination of one immunity group and one vaccination group with the color indicating the percentages of individuals aged 0-4 years in that combination. Summing up the percentages across all small squares within a big square gives 100%. For example, the first big square in Figure S2(a) shows the mapping between immunity and vaccination groups in subpopulation 1 on day 1, with colored small squares indicating that there were individuals aged 0-4 years in the combinations of immunity groups 0-2 (or 5-7) and vaccination groups 0-3.

Based on Figure S2, we see that the history of IPV doses received through essential immunization misclassified high-immunity children as under-vaccinated. For example, at SIAs 3-5, high percentages of individuals were in immunity group 7 but vaccination group 0. Given that the strategy $A^V$-EI tracked only the two IPV doses in Nigeria's essential immunization, individuals fell into vaccination group 0 (unvaccinated) and vaccination groups 1-3 (IPV-only-vaccinated but no more than 2 IPV doses). This explains why there were 0% of individuals in vaccination groups 4-7 under $A^V$-EI.

In contrast the history of OPV2 doses received through oSIAs classified those who acquired sufficient intestinal mucosal immunity through effective nOPV2 doses into high vaccination groups and those received no nOPV2 doses into low vaccination groups. For example, at SIAs 3-5, high percentages of individuals fell into (i) immunity groups 5-7 and vaccination groups 5-7 and (ii) immunity group 0 and vaccination group 0.  Since the strategy $A^V$-oSIA tracked nOPV2 doses in oSIAs, together with the initial condition estimated by the essential immunization records of two IPV doses (see Section **Error! Reference source not found.**), individuals fell into vaccination groups 0-3 and 5-7 (OPV-vaccinated).

(a) Subpopulation 1

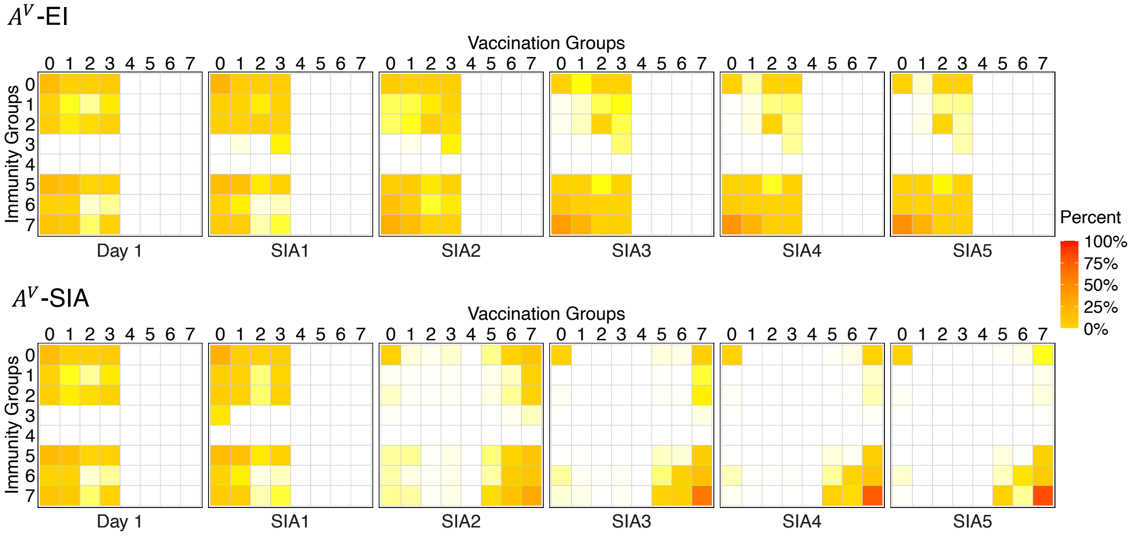


(b) Subpopulation 2

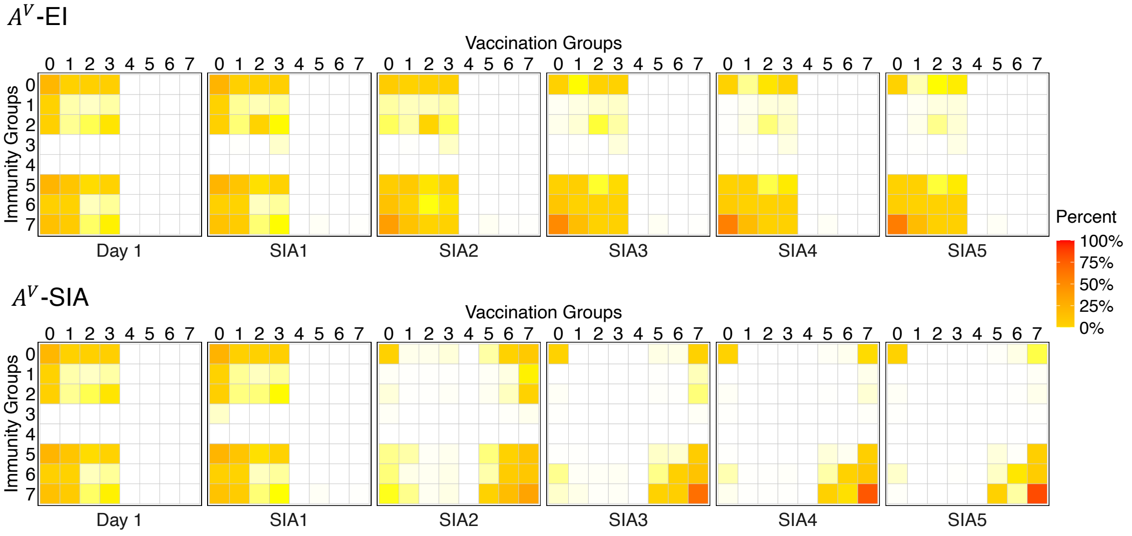


(c) Subpopulation 3

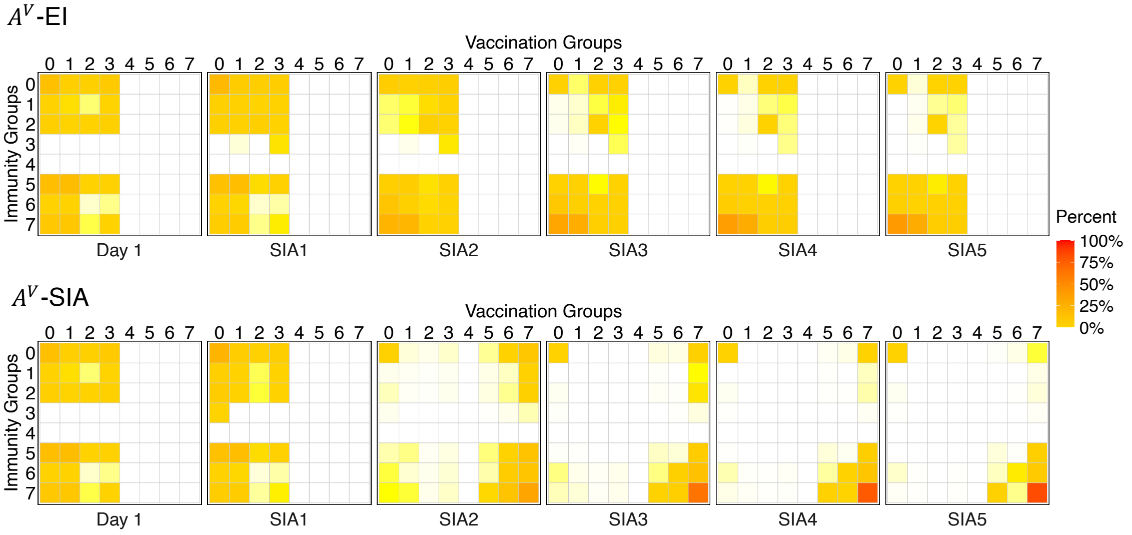

(d) Subpopulation 4

$A^V$-EI

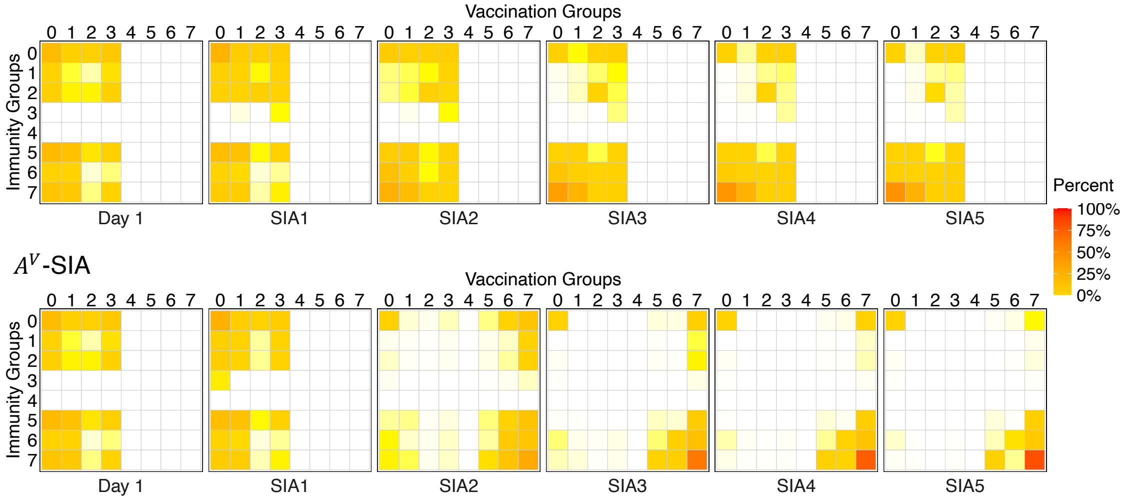


(e) Subpopulation 5

$A^V$-EI

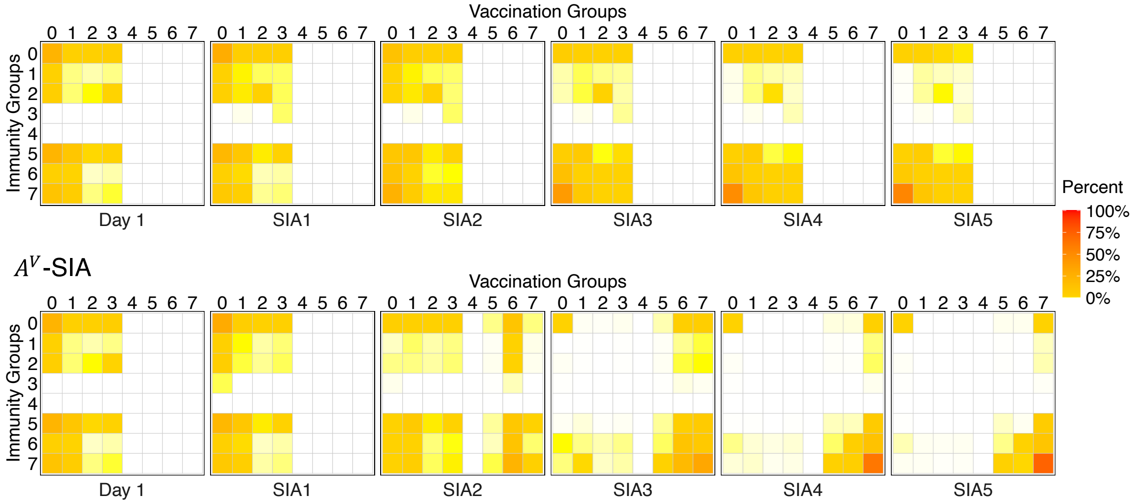


Figure S2: Mapping between immunity and vaccination groups (i.e. the percentage of individuals aged 0-4 years in each combination of an immunity group and a vaccination group) at the start days of simulations (Day 1) and of the 5 subsequent oSIA rounds (SIAs 1-5) in the first outbreak response under vaccine allocation strategies $A^V$-EI and $A^V$-oSIA in (a) subpopulation 1, (b) subpopulation 2, (c) subpopulation 3, (d) subpopulation 4, and (e) subpopulation 5
